# Decreased hospital visits and increased mortality rate in the emergency department during the COVID-19 pandemic: Evidence from Albania

**DOI:** 10.1101/2021.10.07.21264204

**Authors:** Jorgjia Bucaj, Enkeleint A. Mechili, Petros Galanis, Bruna Mersini, Sonila Nika, Inis Hoxhaj, Stefano Likaj, Athina E. Patelarou, Evridiki Patelarou

**Affiliations:** Department of Healthcare, Faculty of Health, University of Vlora, Vlora, Albania; Clinic of Social and Family Medicine, School of Medicine, University of Crete, Crete, Greece; Faculty of Nursing, Center for Health Services Management and Evaluation, National and Kapodistrian University of Athens, Athens, Greece; Vlora Regional Hospital, Vlora, Albania; Department of Nursing, Faculty of Health, University of Vlora, Vlora, Albania; Faculty of Nursing, Hellenic Mediterranean University, Crete, Greece

**Keywords:** COVID-19, emergency department, patient records, hospital visits, mortality rate, SARS-CoV-2, Albania

## Abstract

**Objective:** To investigate the hospital visits and mortality rate during the COVID-19 in emergency department of Vlora regional hospital in Albania and to compare with the three previous years (2017-2019).

**Data sources:** Secondary data of patients that visited emergency department of Vlora Regional hospital Albania (largest hospital in the south of the country), since January 1, 2017 till December 31, 2020.

**Study Design:** This is a retrospective study. We used the hard copy of the patients’ health register records.

**Extraction methods:** The data extraction was conducted during March 2021 till June 2021. Eligible were all patients admitted and recorded in the registry of the Emergency department. The causes of admission were categorized in 14 different disease categories. All registered patient admitted to the Vlora regional hospital were included in the study. Patients that all data were not recorded and patients that data were not possible to be read were excluded.

**Principal Findings:** Study population included 44.917 patients during 2017-2020. Mean age of patients was 51.5 years, while 53.6% were females. The highest number of patients was in 2017 (n=12.407) and the lowest in 2020 (n=9.266). Increase of patients presented with cardiovascular, psychiatric and renal/urinary tract were observed in 2020 in comparison to 2019. Patients decreased over time with an average annual percent decrease of −7% (p-value=0.22). Joinpoint analysis revealed that mortality rate increased over time with an average annual percent increase of 34.3% (95% confidence interval= −42.7% to 214.8%, p-value=0.27).

**Conclusions:** During the study years the number of patients visiting emergency department decreased while mortality rate increased. Educating and raising awareness of patient to seek medical assistance should be a key objective of health policy makers and health personnel. A specific focus should be put on the more vulnerable (elderly and unemployed) as their health status is in higher risk.

## Introduction

Coronavirus disease 2019 (Covid-19) first appeared in Wuhan, China, in December 2019 and quickly spread all over the world.^1^ Till the first week of August 2021, the total number of cases surpassed 200 million, just six months after reaching 100 million. American Region reports the highest number of cases and deaths per 100.000 population followed by Europe.^2^

COVID-19 led to an extensive morbidity and mortality.^3^ Its presentation varies from asymptomatic to severe pneumonia syndrome. Patients with comorbidities such as diabetes, chronic obstructive pulmonary disease (COPD), cardiovascular diseases (CVD), hypertension, malignancies, HIV and others, are more likely to develop a life-threatening situation with higher morbidity and mortality rates.^4^ Specifically, patients with cardiovascular disease, diabetes and hypertension have higher risk for admission to intensive care unit (ICU), mechanical ventilation as well as higher mortality risk.^5^

Hospitals and healthcare systems are facing a catastrophic financial burden due to COVID-19. This situation came as a lack of preparedness and influence patients care and healthcare facilities.^6^ The capacity problem affect the quality of healthcare, making patients more vulnerable to diseases’ complications. In order to add inpatients capacity and staffing, many hospitals are closing out-patients wards and canceling elective procedures.^7^ Coping with the whole situation, healthcare systems will require of public funds to purchase equipment, pay extra staff, and build temporary new wards. There is a global concern that a new post-pandemic period of austerity and long-lasting economic effects will begin.^8^

However, it seems that pandemic has changed general population behaviors, consequently making Emergency Department less visited due to perceived risk of infection.^9^ Admission to emergency department in Finland during the lockdown decreased by 16%.^10^ Additionally, decrease has been reported in Germany, Norway and Greece by 30%, 39% and 42.3% respectively.^11-13^ Additionally, decrease has been reported for different health conditions including cardiovascular^13,14^ and psychiatric^15,16^ issues while for infection diseases^13^ remained stable and higher for respiratory disease.^17^

As all countries, Albania was affected by the COVID-19 pandemic. On March 9, 2020 Albanian Ministry of Health announced the first cases of Covid-19. As of August 14, 2021 have been reported 135.140 new cases and 2.461 deaths in total.^18,19^ In order to deal with the new situation, the government established a task force aiming to trace, prevent and report COVID-19 cases as well as to advise the COVID-19 expert committee. Additionally, the Institute of Public Health prepared guidelines for management of COVID-19 cases.^20^ A crucial role during the management of the whole situation played the calling center of the National Health Emergency Center while provision of healthcare services was adjusted. Initially, the Infectious Disease Services at the University Hospital Centre “Mother Tereza”, (named Hospital COVID-1), and the University Hospital “Shefqet Ndroqi”, (named Hospital COVID-2) provided services to all patients who needed hospitalization. Additionally, all non-urgent medical interventions have been taken to suspend or postponed.^21^ Population (both with chronic and non-chronic conditions) were advised to contact their family doctors by phone for possible services.

However elderly, disabled, homeless and people working in the informal sector were the most affected part of the population, which the state initiated a support program for.^22^ Restriction measures and quarantine had a significant impact on depression levels on Albanian population.^23^ Current state policy objectives are creating protected environment, update information and create response measures, which are seemed being helpful maintaining health indicators constant and decrease pandemic curve.^20^

The COVID-19 pandemic has a significant impact on health status of the population as well as on healthcare system level. This study aimed to investigate the hospital visits and mortality rate during the COVID-19 in emergency department of Vlora regional hospital in Albania and to compare with the three previous years (2017-2019).

## Methods

### Study Design and setting

This retrospective study included patients that visited emergency department of Vlora Regional hospital (Spitali Rajonal i Vlorës), Albania since January 1, 2017 till December 31, 2020. Vlora region is the largest in the south of Albania with a population of around 190.000 people. However in summer the population is growing due to the Albanian emigrants coming back home as well as due to the large number of tourists visiting the city and the seaside places. The hospital also is the largest in south Albania and annually provides services to thousands of people both Albanian and foreigners (especially during summer period). We used the hard copy of the patients’ health registers as electronic databases doesn’t exist yet. The hospital except the emergency department, has different wards including pathology, general surgery-intensive care unit, gynecology – obstetric, otolaryngology-ophthalmology, pediatric, infection diseases unit, microsurgery, anatomopathological - laboratory and oncology-palliative care unite.

### Patients and data extraction

The data extraction was conducted during March 2021 till June 2021. Eligible were all patients admitted and recorded in the registry of the Emergency department of the aforementioned hospital. After ethical approval, the two main researchers (JB and EAM) studied the data in order to decide how to make data extraction and recording of them. The researchers decided to categorize the diseases in 14 different categories. They were cardiovascular (acute myocardial infarction, heart failure, atrial fibrillation and other arrhythmias, heart blocks etc.), neurological (stroke, radicular syndromes, encephalopathy, vertiginous syndromes etc.), pulmonary (asthma, COPD, bronchopneumonia etc.), psychiatric (depression disorders, panic attacks, anxiety etc.), infectious (gastroenteritis, fever syndrome, tonsillitis, insects/reptile bites etc.), gastrointestinal (dyspeptic syndrome, abdominal pain, biliary tract pain, hepatitis, ulcer etc.), renal/urinary tract (renal pain, renal tract infections, urosepsis, renal failure, hematuria etc.), oncological, muscular (lumbar pain, general tiredness, bursitis etc.), toxicological (alcohol intoxication, medicaments intoxication, pesticides intoxication, suicide attempts with different substances, intoxications with narcotic substances), endocrine (mellitus diabetes, thyroid disorders), autoimmune (rheumatoid arthritis, ulcerous colitis, Crohn disease etc.), allergic (hypersensitive reaction from medication/ food/unidentified causes, urticarial, anaphylactic shock etc.) and COVID-19/suspect COVID-19. We decided to include COVID-19 as a specific category despite the fact that especially during the first period of the pandemic tests were conducted by Public Health Department and all patients with severe symptoms were transferred in Tirana. After the first period of the pandemic and when tests were available, emergency unit started making rapid test. Patients with moderate symptoms were admitted at the Vlora hospital in a specific ward that was prepared during summer period.

After admission in the emergency unit, in the patient health record is registered name, surname, age, gender (male / female), social status (employed / unemployed / retired, student), diagnose and outcome (discharge, admitted to hospital, referred to another hospital and death). Laboratory test and other exams conducted weren’t available and we didn’t include them in the study. Regarding traumatic events such as car accidents, gunshot wounds, fractures etc., the admission is made directly in the general surgery-intensive care unit as the Regional Hospital of Vlora doesn’t have a specific emergency unit for traumas. Due to this, these patients were not included in the study. The Emergency department referred in this study is considered as the Internal Diseases Emergency Unit.

After codification of the diseases in the main categories, two different groups started data extraction from the hard copies to excel. The groups (JB/MA and IH/SL) included one physician and one nurse. Both groups used the same codification. In cases that doubts existed communication and discussion between groups was possible.

We did not include in the analysis patients that all data were not recorded (275 patients in total; 102 in 2017; 73 in 2018; 64 in 2019; and 36 in 2020). Additionally, patients that data were not possible to be read from both teams were excluded from the analysis (842 in total; 369 in 2017; 227 in 2018; 116 in 2019; and 130 in 2020). This was mainly due to the non-understandable writing of the doctors or due to the bad condition of the patient health register.

## Statistical analysis

Continuous variables are presented as mean (standard deviation) and categorical variables are presented as numbers (percentages). Age was divided in 10-years intervals in order to calculate mortality by these intervals and gender in hospitalized COVID-19 patients. Differences between year of visit and gender and social status were assessed with chi-square trend test, while differences between year of visit and age were assessed with independent’s sample t-test. In case of t-test, we used Bonferroni post-hoc test to find out differences between subgroups. Also, we used chi-square trend test to compare mortality rate with year of visit and chi-square test to compare mortality rate with gender. These analyses were performed with the Statistical Package for Social Sciences software (IBM Corp. Released 2012. IBM SPSS Statistics for Windows, Version 21.0. Armonk, NY: IBM Corp.).

Moreover, changes in visits in hospitals over time were examined with Joinpoint regression analysis, using Kim’s method^24^ and Joinpoint Regression Program, Version 4.4.0.0 - 2017.^25^ In that case, we evaluated trends over time and we calculated the average annual percent change for visits, the 95% confidence interval and p-values according to gender and age. Age was divided in three categories (<64, 64-80, >80 years) to provide meaningful results. Also, we used Joinpoint regression analysis to identify changes in mortality rate over time. All tests of statistical significance were two-tailed, and p-values <0.05 were considered significant.

## Ethical issues

The study was approved by the Directory of Vlora Regional Hospital with Prot. 4352 Nr. date 28.10.2020 and was signed by the Hospital Director. This Body is responsible for providing ethical approval. The approval was provided based on the memorandum of understanding between University of Vlora “Ismail Qemali” and the Vlora Regional Hospital Nr. 3864 Prot. date 09.10.2015, article 5, point 3 as well as on the Law on Higher Education in Albania Nr. 80/2015 date 22.07.2015, article 93. No personal data were recorded during the data extraction and analysis. The Albanian law Nr.9887, date 10.03.2008 “For Security of Personal data” was strictly followed. All possible data that could identify patients during data analysis or data presentation were excluded. Additionally, ethical principles for the protection of individuals participating in a research study are in line with the Helsinki Declaration for conducting scientific research studies.

## Results

Study population included 44.917 patients during 2017-2020. Demographic and clinical characteristics of patients are shown in Table 1. Mean age of patients was 51.5 years, while 53.6% were females. The highest number of patients was in 2017 (n=12.407) and the lowest in 2020 (n=9.266). Also, most patients were during the summer (n=12.663), and then during the winter (n=11.223), the autumn (n=10.810) and the spring (n=10.221). The most frequent diseases were infection diseases (19.9%), cardiovascular diseases (19.2%), gastrointestinal diseases (18.4%) and neurological diseases (17%). One point four percent of patients visited hospitals due to COVID-19. Out of the 44.917 patients, 112 died (0.2%) and 3.735 were hospitalized (8.3%).

**Table 1.** Demographic and clinical characteristics of patients.

| **Characteristics** | **N** | **%** |
| --- | --- | --- |
| ***Gender*** |  |  |
| Males | 20.825 | 46.4 |
| Females | 24.079 | 53.6 |
| ***Age, mean, standard deviation*** | 51.5 | 20.7 |
| ***Year*** |  |  |
| 2017 | 12.407 | 27.6 |
| 2018 | 11.328 | 25.2 |
| 2019 | 11.916 | 26.5 |
| 2020 | 9.266 | 20.6 |
| ***Month*** |  |  |
| January | 4.092 | 9.1 |
| February | 3.896 | 8.7 |
| March | 3.435 | 7.6 |
| April | 3.170 | 7.1 |
| May | 3.616 | 8.1 |
| June | 4.191 | 9.3 |
| July | 4.383 | 9.8 |
| August | 4.089 | 9.1 |
| September | 4.100 | 9.1 |
| October | 3.464 | 7.7 |
| November | 3.246 | 7.2 |
| December | 3.235 | 7.2 |
| ***Social status*** |  |  |
| Employed | 9.498 | 22.8 |
| Unemployed | 13.555 | 32.6 |
| Retired | 16.233 | 36.1 |
| Students | 2.293 | 5.5 |
| ***Disease/disorder*** |  |  |
| Cardiovascular | 8.538 | 19.2 |
| Neurological | 7.565 | 17.0 |
| Pulmonary | 1.556 | 3.5 |
| Psychiatric | 1.174 | 2.6 |
| Infection | 8.880 | 19.9 |
| Gastrointestinal | 8.198 | 18.4 |
| Renal/urinary tract | 2.815 | 6.3 |
| Oncological | 128 | 0.3 |
| Muscular | 393 | 0.9 |
| Toxicological | 2.050 | 4.6 |
| Endocrine | 496 | 1.1 |
| Autoimmune | 221 | 0.5 |
| Allergic | 896 | 2.0 |
| COVID-19 | 626 | 1.4 |
| ***Outcome*** |  |  |
| Discharged | 41.019 | 91.4 |
| Hospitalized | 3.735 | 8.3 |
| Died | 112 | 0.2 |

Increase of patients presented with cardiovascular (18.7% to 22.2%), psychiatric (0.4% to 3.4%) and renal/urinary tract (5% to. 6.8%) were observed in 2020 in comparison to 2019 (Table 2). In contrast, a significant decrease was observed for other diseases the same period (neurological 17.1% to 13.7%, pulmonary 3.6% to 2.9%, infection 21.5% to 17.2%, gastrointestinal 21.6% to 16%). Additionally, for some disease, prevalence remained stabled or small changed were observed (muscular 1.1% to 1.4%, toxicological 4.2% to 4.3% etc.).

**Table 2:** Prevalence of reported diseases and outcome 2017-2020

|  | Year |  |  |  |  |  |  |  |
| --- | --- | --- | --- | --- | --- | --- | --- | --- |
|  | 2017 |  | 2018 |  | 2019 |  | 2020 |  |
|  | N | % | N | % | N | % | N | % |
| Disease/disorder |  |  |  |  |  |  |  |  |
| Cardiovascular | 2383 | 19.2 | 1930 | 17.1 | 2175 | 18.7 | 2050 | 22.2 |
| Neurological | 2224 | 18.0 | 2090 | 18.5 | 1989 | 17.1 | 1262 | 13.7 |
| Pulmonary | 443 | 3.6 | 427 | 3.8 | 422 | 3.6 | 264 | 2.9 |
| Psychiatric | 521 | 4.2 | 295 | 2.6 | 48 | 0.4 | 310 | 3.4 |
| Infection | 2314 | 18.7 | 2481 | 21.9 | 2498 | 21.5 | 1587 | 17.2 |
| Gastrointestinal | 2229 | 18.0 | 1981 | 17.5 | 2508 | 21.6 | 1480 | 16.0 |
| Renal/urinary tract | 914 | 7.4 | 690 | 6.1 | 584 | 5.0 | 627 | 6.8 |
| Oncological | 38 | 0.3 | 56 | 0.5 | 3 | 0.0 | 31 | 0.3 |
| Muscular | 113 | 0.9 | 11 | 0.2 | 131 | 1.1 | 127 | 1.4 |
| Toxicological | 612 | 4.9 | 551 | 4.9 | 490 | 4.2 | 397 | 4.3 |
| Endocrine | 101 | 0.8 | 112 | 1.0 | 207 | 1.8 | 76 | 0.8 |
| Autoimmune | 36 | 0.3 | 163 | 1.4 | 7 | 0.1 | 15 | 0.2 |
| Allergic | 242 | 2.0 | 248 | 2.2 | 216 | 1.9 | 190 | 2.1 |
| COVID-19 | 0 | 0 | 0 | 0 | 0 | 0 | 626 | 6.8 |
| Others | 219 | 1.8 | 268 | 2.4 | 329 | 2.8 | 190 | 2.1 |
| Outcome |  |  |  |  |  |  |  |  |
| Discharged | 11,401 | 91.9 | 10,390 | 91.7 | 10,901 | 93.0 | 8,327 | 88.5 |
| Hospitalized | 980 | 7.9 | 915 | 8.1 | 805 | 6.9 | 1035 | 11.0 |
| Died | 26 | 0.2 | 23 | 0.2 | 17 | 0.1 | 46 | 0.5 |

Joinpoint analysis of the patients visited hospitals during 2017-2020 according to gender and age is shown in Table 3. Patients decreased over time with an average annual percent decrease of −7% (p-value=0.22). Also, males and females patients decreased over time with an average annual percent decrease of −4.5% (p-value=0.34) and −9% (p-value=0.16) respectively. A similar decreased trend was found in patients ≤80 years, but the opposite was found in patients >80 years. In particular, patients >80 years increased over time with an average annual percent increase of 1.6% (p-value=0.87).

**Table 3.** Joinpoint analysis of the patients visited hospitals during 2017-2020 according to gender and age.

|  | **Average annual**<br>**percent change** | **95% confidence**<br>**interval** | **P-value** |
| --- | --- | --- | --- |
| **Total** | -7.0 | -22.1 to 11.1 | 0.22 |
| **Gender** |  |  |  |
| Male | -4.5 | 18.8 to 12.3 | 0.34 |
| Female | -9.0 | -25.0 to 10.3 | 0.16 |
| **Age** |  |  |  |
| <64 years | -8.7 | -24.0 to 9.7 | 0.16 |
| 64-80 years | -4.8 | -16.2 to 8.2 | 0.24 |
| >80 years | 1.6 | -29.0 to 45.5 | 0.87 |

During 2017-2020 females visited more often hospitals than males (53.6% vs. 46.4, x^2^=48, p-value for trend<0.001). Also, retired (39%) and unemployed (32.6%) visited more often hospitals than employed (22.8%) and students (5.5%), (x^2^=105, p-value<0.001). Mean age of patients in 2020 (52.8 years) was higher than 2019 (51.9 years, t=3.2, p-value=0.009), 2018 (51.7 years, t=3.9, p-value=0.001), and 2017 (50.2 years, t=9, p-value<0.001).

Joinpoint analysis revealed that mortality rate increased over time with an average annual percent increase of 34.3% (95% confidence interval= −42.7% to 214.8%, p-value=0.27). In particular, 0.5% of patients in 2020 died, while the respective percentages in 2017, 2018, and 2019 were 0.2%, 0.2%, and 0.1%. Mortality rate was higher among >80 years (0.6%) and 64-80 years (0.5%) than <64 years (0.1%), (x^2^=700, p-value<0.001). Also, males died more often than females (0.3% vs. 0.2%, x^2^=117, p-value<0.001).

## Discussion

This study aimed to investigate the hospital visits and mortality rate during the COVID-19 pandemic in emergency department of Vlora regional hospital in Albania. During the study years (2017-2020), the number of patients visiting emergency department decreased. The lowest number of visits was observed during the 2020, the year of the COVID-19 pandemic. The number of patients with cardiovascular, psychiatric and renal/urinary tract increased in 2020 in comparison to 2019 while those with neurological, pulmonary, infection and gastrointestinal diseases decreased during the same period. Additionally, mortality rate was stable for 2017-2019 but increased significantly in 2020. A first analysis revealed that during the COVID-19 pandemic in emergency department the number of visiting patients decreased while the mortality rate increased.

A study conducted in USA reported that number of visits in the emergency department was significant lower during the COVID-19 period in comparison to pre-COVID-19 pandemic.^26^ Another US study revealed that during the first period of the pandemic, the number of patients visiting emergency department was 42% lower than a year before.^27^ Similar results with the current work have been reported also in other studies conducted in Europe and elsewhere for both pediatric and non-pediatric population.^10,15,28-30^ According to Albanian Institute of Statistics (INSTAT) the number of hospitalized person has increased annually during the period 2017-2019. ^31^ However, data are not available yet for 2020. Despite the low number of infections with COVID-19 in Albania during the first phase of the pandemic, we saw a decreased number of patients visiting emergency ward. The lower number of visits in emergency department during the COVID-19 period can be explained due to different reasons. The implementation of the lockdown, restrictions in movement and closure of non-essential activities most probably contributed to this reduction. The fear of getting infected with COVID-19 most likely kept many person out of visiting the emergency ward despite their possible health problem. Additionally, the advice from Albanian health authorities to stay home and to visit hospitals only for urgent issues and after calling at the National Health Emergency Center may be another possible explanation. Furthermore, population was advised to keep close contacts with their family doctors for their health conditions. Another possible explanation for this decrease can be the fact that many visits to the emergency department before the COVID-19 pandemic were not-necessary and not urgent. However, more data and studies are needed in order to accept or not this explanation. To some extent, majority of patients visiting emergency ward are reported in summer. During 2020, border closure didn’t permit to Albanians living abroad and tourist to visit the country. This most probably has also contributed to the low number of admissions in emergency department in 2020. An extensive analysis about patients that had immediate and urgent life threating needs could help in better understanding this drop. Additionally, policymakers jointly with health professionals and patient associations should work in order to increase awareness for asking medical assistance. This is very crucial not only now during the COVID-19 period but in general as there is a lack of prevention culture among Albanians^32^ and they usually ask for medical assistance in later stages of a disease.

The current study reported an increase in mortality rate during 2020 while the previous years was stable or even lower for 2019. According to the annual report of INSTAT, death toll increased by 25.8% in 2020 in comparison to 2019.^33^ According to the same report, death rates of male population is higher than those of female. Main death causes by the report are diseases of the circulatory system while in the current study the number of cardiovascular patients visiting emergency department is reported second after infectious diseases. To some extent, the main factor of death in Vlora city are diseases of the circulatory system.^33^

This higher mortality in 2020 in comparison to the previous year most probably is connected with the ongoing pandemic. Data in Albania show that as about 31 December 2020, there were diagnosed 58.316 new cases of COVID-19 and 1181 deaths in total.^34^ Due to all causes, in Albania in 2020, there were reported in total 27.605 fatalities.^33^ Most probably the increase of death toll is connected with the COVID-19 situation. The higher number is not only due to the new cause of death but also due to late seek of medical assistance. Patients due to the fear of getting infected with COVID-19, postponed their visits to hospital. Most probably, this phenomena aggregated their health status and lead to death. This has been reported as *“collateral damage”* due to the pandemic by Marsoor (2020).^35^ Mortality rate after COVID-19 was doubled in emergency department in Lebanon in comparison to pre-Covid-19 period.^36^ A study in northern Italy reported lower admission rates in emergency department as well as an increase in out-of-hospital mortality for all causes.^37^

The mean age of patients presented in emergency ward after COVID-19 period was higher than before the pandemic. Similar results are presented also in another study.^36^ In general, COVID-19 disease severity and mortality is higher for older people. Fear of getting infected and a possible negative outcome most probably made them visit more often the emergency department.

Increase of visits to emergency department was reported for cardiovascular, psychiatric and renal/urinary tract and decrease for neurological, pulmonary, infection and gastrointestinal diseases. Decrease of admissions to emergency department for cardiovascular diseases was observed in different studies.^13,38^ Psychiatric events have been reported higher during the COVID-19 period in Kentucky^39^ while in other studies the opposite was reported.^15,40^ These heterogeneity may be due to the differences in restriction measures undertaken by different governments. However, studies in Albania have reported a high prevalence of mental health issues during the quarantine period.^23,41^ Decrease of infection diseases events has been also reported in other studies.^10^ This decrease is mainly due to the lockdown and the restriction measures. The lower rates of seasonal influenza in 2020 due to the restriction measures and the prevention actions undertaken, most probably contributed to lower admission rates.

## Strengths and limitations

To our best knowledge, this is the first study in Albania that aimed to examine the hospital visits and mortality rate during the COVID-19 in emergency department of Vlora regional hospital in Albania. In order to have a better overview of the situation we compared 2020 admissions with the three previous years (2017-2019) and not only with several months or just the previous years as many other studies have done. The high number of data analyzed is among the biggest strength of the current study. Additionally, analysis of different diseases make clearer the actual situation and the real impact of the pandemic in admission to emergency ward. The use of the Joinpoint analysis is also a pro of the current study.

However, as all studies, the current work suffers from some limitations. Including in the analysis data from only one regional hospital make difficult generalization. Additionally, as data were retrieved from hard copy patients registers, difficulties were found in reading the document due to non-understandable writing and documents decay.

## Conclusions

The current study aimed to assess hospital visits and mortality rate during the COVID-19 period in emergency department in Albania. Number of patients that visited emergency department decreased in 2020 in comparison to previous years (2017-2019). Infection, cardiovascular and gastrointestinal diseases were the most common causes of visits. Additionally, 1,4% visited the ward due to COVID-19 infection. During the study period, females visited more often hospitals than males while mean age of patients in 2020 was higher than the three previous years. Mortality rate increased over time with an average annual percent increase of 34.3%. In particular, 0.5% of patients in 2020 died, while the respective percentages in 2017, 2018, and 2019 were 0.2%, 0.2%, and 0.1%. Future studies should include more hospitals in their analysis as well as should focus on the reasons of admission drop. Additionally, is of paramount significance to study which patient had immediate and urgent life threating needs and visited the hospital and who of them not. A specific focus should be put on the reasons of deaths during the 2020 and how many deaths could be avoid if earlier medical assistant was asked. Educating and raising awareness of patient to seek medical assistance should be a key objective of health policy makers and health personnel. To some extent, more attention during services provision should be given to the more vulnerable (elderly and unemployed) as their health status is in higher risk. However, continuation and improvement of virtual health services provision could ameliorate the situation. Finally, development of mobile health teams (physician and nurse) for home visits is strongly recommended.

## Data Availability

Data are available after reasonable request.

## Acknowledgements

Authors would like to thank Mateo Alushi for his assistance.

